# Minimum required data for decision at a revision MDT in an arthroplasty network: a structured protocol for a systematic review of the literature and consensus statement using a two round Delphi process

**DOI:** 10.1101/2025.10.14.25337994

**Authors:** J Shelton, S Dorman

## Abstract

Total hip arthroplasty is one of the most successful operations of all time however the British NJR demonstrates a revision burden of 1.8% at 13.5 years. This creates a burden of 1500 revisions per annum across the UK. Various service improvement projects have demonstrated a need to consolidate specialist surgeries in specialist centres mandating the formation of revision networks. Exactly what information is required from a referring centre to make an on the day decision at a regional MDT has not been assessed.

We aim to build a consensus on a minimum required data set for referral into a regional arthroplasty network in order to allow said MDT to categorise the revision indication, complexity and prescribe a bespoke surgical plan for the revision.

A literature search will be performed across medical search engines to identify abstracts related to surgical decision making in revision arthroplasty. All abstracts will be assessed by two assessors and included or rejected for full paper retrieval. Every input for decision making within the included papers will be recorded.

Once the input set is completed a group of experts in hip surgery will be invited to participate in a two round Delphi process to stratify the importance of each input. Once this process is completed a consensus setting meeting of all stakeholders will be held to formalise a consensus.

It is anticipated that the consensus statement from this process will be used as a framework for regional if not nation-wide revision network referral forms.

## Background

Total hip arthroplasty is one of the most successful operations receiving the accolade of operation of the century in the Lancet [1]. Recent data from the NJR states the risk of revision at 13.5 years is 1.8%. This will lead to and additional 1500 joints requiring revision per annum [2].

Since 2015 a number of high profile reports such as Getting It Right First Time [3] and the BOAST standard for revision knee surgery [4] have been released through professional societies making the case for revision services to be consolidated into units with appropriate critical mass [5]. This in turn has led to the formation of revision networks allowing surgeons in a central unit to ensure the meeting the critical mass of cases to maintain skill in continuous professional development in revision arthroplasty.

The new working model should include the discussion of all revision cases with a central Major Revision Network (MRC) through a combined MDT between all the units within the network. The issue arises of exactly what information is required to allow decision making at the MRC MDT when referring from a primary or revision unit? There is a paucity on data regarding the clinical assessment or tests required for effective decision making. An emerging strategy to overcome this type of issue is the development of core outcome sets [6] [7] [8] [9] [10] [11] [12] [13] [14] [15] [16]. These provide a minimum expected data set for sites to collect to allow effective meta-analysis or randomised control trials across numerous networks. In recent years several core outcome sets have been introduced across orthopaedics [17] [18] [19]. Core outcome sets provide and excellent framework for identification of key parameters and ranking their importance across multiple stakeholder groups. Using the same framework adapted from Dorman et al. in trials 2018 would allow for the development of a core input set for information required for MDT decision making in revision surgery through a systematic review and two round Delphi process for both hip and knee revision. .

## Aims & Objective

### Aim

The aim of this study is the development of a consensus on the data required for both practical decisions making during regional MDT sessions with the secondary aim of use for observational research and clinical trials on revision hip & knee surgery.

### Objectives

#### The specific study objectives are

- To identify reported diagnostic tests, clinical measurements or patient reported outcome measures denoted in randomised control trials (RCTs), case-control studies, case series from a systematic review of the literature.
- To prioritise these outcomes of primary surgery including diagnostic tests, clinical measurements or patient reported outcome measures from the perspective of key stakeholder groups using a two-round Delphi
- To conduct a consensus meeting, compare all diagnostic tests, clinical measurements and patient reported outcome measures considered important to all stakeholders and to integrate the required pre-MDT information into a combined core outcome set.

### Methods/Design

This methodology has been adapted from Dorman et al. published in Trials in 2018. This project has been registered with Core Outcome Measures in Effectiveness Trials (COMET).

### Search strategy

A comprehensive literature search was conducted with the assistance of an information specialist from the University of Sheffield. The following databases were searched from 1 January 1990 to 23 November 2022: MEDLINE (Ovid), Scopus, and the Cochrane Central Register of Controlled Trials (CENTRAL). An arbitrary cut-off of 1990 was selected to capture contemporary practice. No restrictions were applied with regard to study design, language, or publication status.

Full electronic search strategies for each database, including all keywords and MeSH terms used, are provided in Appendix 1. In brief, the strategies combined controlled vocabulary and free-text terms for revision joint replacement surgery and arthroplasty, in conjunction with terms relating to decision making, shared decision making, multidisciplinary teams (MDTs), and patient care teams.

### Eligibility criteria

#### Studies were eligible for inclusion if they

- Reported outcomes relating to revision joint replacement or reoperation following hip or knee arthroplasty.
- Included reference to decision-making processes, multidisciplinary team (MDT) input, clinical measures, diagnostic tests, or patient-reported outcome measures (PROMs).
- Were published between 1990 and 2022.

#### Exclusion criteria were

- Non-human studies.
- Narrative reviews, editorials, commentaries, or conference abstracts without primary data.
- Studies not reporting outcomes relevant to pre-MDT decision making.

### Study selection

All titles and abstracts identified through the database search were imported into Rayyan for screening. Two reviewers independently screened all records. Any titles or abstracts considered potentially eligible were retrieved for full-text review. Full-text screening was also performed independently by two reviewers, with disagreements resolved through discussion. Where consensus could not be reached, a third reviewer adjudicated.

### Data extraction

Data from included studies will be extracted into a standardised form designed for this review. Extracted data will include bibliographic details, study design, patient population, type of revision surgery, decision-making processes described, clinical outcomes, diagnostic measures, and PROMs. No data synthesis or meta-analysis is planned, as the purpose of this review is to generate a comprehensive list of relevant outcomes for subsequent use in a Delphi process.

### Risk of bias assessment

Given the descriptive objective of the review, no formal risk of bias or quality assessment will be undertaken. This approach was chosen as the review is intended to identify outcome measures rather than to synthesise evidence of effectiveness.

### Reporting

The review will be reported in accordance with the Preferred Reporting Items for Systematic Reviews and Meta-Analyses (PRISMA) guidelines. A PRISMA flow diagram will be used to document the process of study identification, screening, eligibility assessment, and inclusion.

### Outcomes

The primary aim of this project is to identify all pre-op or pre-MDT diagnostic tests, clinical measurements or patient reported outcome measurements which may aid in diagnosis and proposed management plan to be refined by a Delphi Process to a minimum expected information core outcome set for referral to the network MDT.

### Data analysis & Presentation

This project will use the comprehensive framework of health to develop a core outcome set, this favours the validity of the product. Each parameter will be assigned to one of the five core areas described within the Dodd-Williamson classification. These include (1) Adverse Events, (2) Death, (3) Physiological/clinical, (4) Life impact and (5) Resource use. A sixth domain may be added for included measurements that do not fit elsewhere within the framework [20].

Within each domain we will evaluate the number of measurements used, time points pre-MDT in which they were measured and method of measurement.

### Identification of potential measurements

A list of all potential measurements will be identified in the course of this systematic review. These measurements will be listed both individually and by domain. All measurements will be reviewed by the study steering group (SSG). The study steering group will consist of the authors along with representative from British hip society and British association of knee surgery. Representatives from patient advocacy groups will also be invited to provide input on patient reported outcome measures and clinical measurements.

### Identification of importance of measurements

#### Overview

The importance of each identified measurement will be assessed by a group of experts using a two round Delphi process.

#### Participants

The Delphi survey will be conducted using a group of clinicians with a special interest in revision hip and knee surgery through the specialist societies BHS and BASK. Clinicians will only be invited to participate if they are currently involved with revision hip and or knee surgery.

Clinician leaders will be identified through the British Hip Society (BHS) and British Association of Surgeons of the Knee (BASK). Eligible participants will be contacted via e-mail and asked to complete an online Delphi questionnaire. A minimum of 30 surgeons will be recruited for this process.

The number of participants will be noted at each stage from agreeing to be part of the process, round 1 and round 2 of the Delphi process.

Bespoke Delphi management software will be used to ensure information if recorded against a participant identifier.

#### Delphi Survey

##### Delphi Round 1

The first-round questionnaire will be used to collect demographic information on the Delphi participants including name, clinical role, place of work, e-mail address. Personal information will be stored in a separate database with a unique identifier.

At each round participants will be allocated 3 weeks to complete the survey with a reminder at the end of week 2. Participants who do not complete round 1 will not be invited to complete round 2 and will be excluded from the study.

All data will be collected using online surveys. A list of measurements will be scored on a Licat scale of importance and will have an option to add additional measurements during round 1. The Grading of Recommendations, Assessments and Evaluations scale will be used to score the measurements. Gradings will be possible from 1-9 with 1-3 deemed not important, 4-6 important but not critical and 7-9 deemed critical importance.

##### Analysis of round 1

All additional measurements suggested by participants to be reviewed by the two reviewers to ensure they do not represent repeated measurements. If any doubt a third assessor will be utilised.

The number of assessors that scored each measurement will be recorded and the distribution of the scores summarised. All outcomes will be taken forward to round 2.

##### Response rate round 1

The response rate will be assessed and presented as a percentage of the total number of participants. Continuation to round 2 will depend on the response rate of round 1. In the event of low response rate (<10) the protocol for future rounds will be reviewed. Where the responses are homogenous the SSG may include without a second round.

The data from round 1 will be presented to the participants and the participants asked to rescore the measurements in round 2. Participants will then be asked to re-score the measurements after review of the round 1 data and distribution.

On completion of round 2 all measurements will be analysed for inclusion into a core outcome set – 1-3 consensus out, 4-6 no consensus and 7-9 consensus in. Additional round of the Delphi process could be required to achieve consensus and will be considered by the SSG on completion of round 2.

### Consensus meeting

The final exercise will be a consensus setting exercise consisting of a consensus focus group and the SSG. Round 2 results will be presented, discussed and voting performed for final inclusion into the core outcome set.

### Definition of consensus

The classification of consensus is outlined in table 3 and will be used to determine if consensus has been achieved.

**Table 2.** Overview of modified Dodd-Williamson [21] classification of outcomes.

| Core area | Core domains | Example |
| --- | --- | --- |
| Adverse events | Adverse events | Unintended consequences |
| Death | N/A | N/A |
| Physiological/clinical | Musculoskeletal | Mal-union, non-union, range of motion |
| Life impact | Physical/social/role/emotional/cognitive functioning<br>Health-related quality of life (HRQL)<br>Delivery of care* | PROMS, activities of daily living, satisfaction |
| Resource use | Economic/hospital/need for intervention, societal burden | Length of stay, further surgery, physiotherapy |
| Technical considerations | Technical/surgical considerations | Radiographic measurements |

\*Delivery of care does not refer to the resource delivery, but instead includes patient satisfaction, patient preference, adherence, withdrawal, tolerability; PROMS, patient-reported quality of life, N/A not applicable

**Table 3.** Classification of consensus[22].

| Consensus classification | Description | Definition |
| --- | --- | --- |
| Consensus in | Consensus that outcome should be included in the core outcome set | 70% or more participants scoring as 7 to 9 *and* < 15% participants scoring as 1 to 3 |
| Consensus out | Consensus that outcome should not be included in the core outcomes set | 70% or more participants scoring as 1 to 3 *and* < 15% of participants scoring as 7 to 9 |
| No consensus | Uncertainty about importance of outcome | Anything else |

Consensus included will have been achieved if a large majority (>70%) score a measurement with 7-9 indicating the measurement is of critical importance and with a small minority (<15%) finding it of no clinical importance (1-3). The reverse is also true, consensus excluded will be achieved if the vast majority score a measurement of no clinical importance (1-3) and only a small minority score the measurement of critical importance (<15%) [21]

In the event of no consensus (4-6) the consensus focus group will make the final decision on the measurement through discussion of opinions and final decision using the Nominal Group Technique (NGT).

## Discussion

There is currently no published minimum required data set of decision in revision arthroplasty. With the implementation of the revision arthroplasty networks encouraged by GIRFT, BHS, BASK and the BOA we as revision surgeons have a golden opportunity to develop a core outcome set to ensure all networks have safe and effective decision making but also allowing meta-analysis of the input and output of networks to build collaboration and research in this challenging area of orthopaedic practice.

## Supporting information

Appendix 1 - Search terms

## Data Availability

Appendix 1 denotes the search terms for the systematic review for reproducibility

