## Appendix 1 - Search terms for "Minimum required data for decision at a revision MDT in an arthroplasty network: a structured protocol for a systematic review of the literature and consensus statement using a two round Delphi process"

**Appendix 1. Full Electronic Search Strategies**

**Database:** Ovid MEDLINE(R) and Epub Ahead of Print, In-Process, In-Data-Review & Other Non-Indexed Citations and Daily
**Coverage:** 1946 to 23 November 2022
**Date searched:** 23 November 2022
**Hits:** 239

1. exp Arthroplasty, Replacement/

2. Reoperation/

3. 1 and 2

4. (revision adj3 (joint replacement* or arthroplast* or hip replacement* or knee replacement* or knee surgery or hip surgery)).ti,ab.

5. 3 or 4

6. decision making/ or decision making, shared/

7. (decision making or MDT or multidisciplinary team).ti,ab.

8. Patient Care Team/

9. 6 or 7 or 8

10. 5 and 9

**Database:** Scopus
**Date searched:** 23 November 2022
**Hits:** 124

( ( TITLE ( revision W/3 ( "joint replacement*" OR arthroplast* OR "hip replacement*" OR "knee replacement*" OR "knee surgery" OR "hip surgery" ) )

OR ABS ( revision W/3 ( "joint replacement*" OR arthroplast* OR "hip replacement*" OR "knee replacement*" OR "knee surgery" OR "hip surgery" ) ) ) )

AND

( ( TITLE ( ( "decision making" OR mdt OR "multidisciplinary team" ) )

OR ABS ( ( "decision making" OR mdt OR "multidisciplinary team" ) ) ) )

**Database:** Cochrane Library (CENTRAL – Cochrane Central Register of Controlled Trials)
**Date searched:** 23 November 2022
**Hits:** 5

#1 MeSH descriptor: [Arthroplasty, Replacement] explode all trees

#2 MeSH descriptor: [Reoperation] explode all trees

#3 #1 AND #2

#4 (revision near/3 ("joint replacement*" or arthroplast* or "hip replacement*" or "knee replacement*" or "knee surgery" or "hip surgery")):ti,ab,kw

(Word variations have been searched)

#5 #3 OR #4

#6 MeSH descriptor: [Decision Making] this term only

#7 MeSH descriptor: [Decision Making, Shared] this term only

#8 ("decision making" or decision-making or MDT or "multidisciplinary team"):ti,ab,kw

(Word variations have been searched)

#9 MeSH descriptor: [Patient Care Team] this term only

#10 #6 or #7 or #8 or #9

#11 #5 and #10
